# Periodontal inflammation mediates the link between homocysteine and high blood pressure

**DOI:** 10.1101/2021.02.02.21251010

**Authors:** João Botelho, Vanessa Machado, Yago Leira, Luís Proença, José João Mendes

**Affiliations:** Clinical Research Unit (CRU), Centro de Investigação Interdisciplinar Egas Moniz (CiiEM), Egas Moniz – Cooperativa de Ensino Superior, Almada, Portugal; Evidenced-Based Hub, CiiEM, Egas Moniz – Cooperativa de Ensino Superior, Almada, Portugal; Periodontology Unit, UCL Eastman Dental Institute and NIHR UCLH Biomedical Research Centre, University College London, London, UK; Periodontology Unit, Faculty of Medicine and Odontology, University of Santiago de Compostela; Medical-Surgical Dentistry (OMEQUI) Research Group, Health Research Institute of Santiago de Compostela (IDIS), Santiago de Compostela, Spain; Clinical Neurosciences Research Laboratory, Health Research Institute of Santiago de Compostela (IDIS), Santiago de Compostela, Spain; Quantitative Methods for Health Research (MQIS), CiiEM, Egas Moniz – Cooperativa de Ensino Superior, Almada, Portugal

**Keywords:** NHANES, periodontal diseases, periodontitis, homocysteine, inflammation, blood pressure, oral health

## Abstract

**Background:** Here, we assess the association between Hcy serum levels and periodontal status in a large representative sample of the National Health and Nutrition Examination Survey (NHANES).

**Methods:** We included 4,021 participants with a periodontal examination, medical self-reported data, BP and blood samples to determine complete blood count, C-reactive protein (CRP) and Hcy levels. We then calculated the periodontal inflamed surface area (PISA) and the periodontal epithelial surface area (PESA). Multivariable regression analysis explored the association between Hcy, periodontal measures and blood pressure. Mediation analysis was performed to understand the effect of age on the association of periodontitis with BP. Mediation analysis assessed the effect of PISA and PESA in the link between Hcy and BP.

**Results:** 4,021 participants fulfilled the inclusion criteria. Hcy levels showed significant correlations with systolic BP, diastolic BP, PISA, PESA and age. PESA showed to be significantly associated with Hcy both for the crude and adjusted models (p<0.01), but not PISA (p>0.05). In the association of Hcy with systolic BP, PISA significantly mediated 17.4% and PESA 0.9%. In the association of Hcy with diastolic BP, PISA significantly mediated 16.3% and PESA 47.2%.

**Conclusions:** This report shows that Hcy and periodontitis are associated. Also, both PISA and PESA significantly mediated the association of Hcy with systolic BP and diastolic BP. Future studies shall deepen the mechanisms by which homocysteine levels increase in a clinical situation of periodontitis.

**One-sentence summary describing the key finding(s) from the study:** Homocysteine and periodontitis are associated. The periodontal inflamed and epithelial surface areas significantly mediate the association of homocysteine with systolic and diastolic blood pressures.

## Introduction

Homocysteine (Hcy) is an amino acid synthesized as an intermediate metabolite from the methionine (Met) biosynthesis ^1,2^. In plasma, 1% of Hcy circulates in its free form while the remaining 99% is bound to proteins ^3^, and its concentration normally ranges between 5-15 µmol/L ^2^. In cases of hyperhomocysteinemia (defined as total Hcy in plasma >15 µmol/L) ^4,5^, the main causes may be impaired metabolism of Met or defective cofactors in the pathways of vitamins B6, B12 and folic acid ^6^.

Hyperhomocysteinemia has been associated with several systemic conditions ^7–16^. In particular, Hcy levels are significantly associated with high blood pressure ^17^ as they inhibit the synthesis of endothelial nitric oxide causing its dysfunction and damages the myocardium via the excessive production of reactive oxygen radicals ^18,19^. Hcy was firstly introduced as a candidate in the link of periodontal and cardiovascular conditions ^20^, but ever since studies on this topic have been relatively scarce.

Periodontitis is a chronic inflammatory condition of the periodontium driven by a dysbiotic plaque ^21,22^. Hcy was firstly reported to be associated with periodontitis ^23,24^, and was suggested as an inflammatory marker of periodontitis as its levels decrease after periodontal treatment ^25^. Moreover, periodontitis plays a very active role in cardiovascular illnesses (for instance, high blood pressure) ^26^, and here a possible form of action of periodontitis emerges in the Hcy-hypertension link (i.e., mediation effect), however this has never been studied.

The primary aim of the present study was to assess the association between Hcy serum levels and periodontal status in a large representative sample of the National Health and Nutrition Examination Survey (NHANES). As secondary aim, we appraised the mediation effect of periodontal measurements in the link between Hcy and blood pressure (BP).

## Methods

### Study Design and Participants

In this study, we performed a secondary analysis using datasets from the NHANES of 2001-2002 and 2003-2004. Data was gathered through interviews, physical and laboratory exams, and both waves have been reviewed and approved by the Centers for Disease Control (CDC) and Prevention National Center for Health Statistics Research (NCHS) Ethics Review Board. All included participants provided a written informed consent.

For the purpose of this study, the following inclusion criteria were defined: participants with 18 years old or older, undergone periodontal examination and had completed measurement of plasma Hcy. This study follows the STrengthening the Reporting of OBservational studies in Epidemiology (STROBE) guideline ^27^.

### Periodontal examination

The periodontal examination was carried out using a partial-mouth protocol using randomly assigned quadrants (one on the upper arch, and one on the lower arch), by calibrated examiners as previously defined ^28^. Three periodontal measures were registered: probing pocket depth (PPD), clinical attachment loss (CAL) and bleeding on probing (BOP). These measures were made at three buccal sites per tooth (mesio-, mid- and disto-buccal). The presence of PD was diagnosed according to the CDC/American Academy of Periodontology case definition (Page & Eke 2012). Then, PD was categorized as mild (≥2 interproximal sites with CAL ≥3 mm; and ≥2 interproximal sites with PPD ≥4 mm [not on the same tooth] or 1 site with PPD ≥5 mm), moderate (≥2 interproximal sites with CAL ≥4 mm [not on the same tooth] or ≥2 interproximal sites with PPD ≥5 mm, also not on the same tooth) and severe (≥2 interproximal sites with CAL ≥6 mm [not on the same tooth] and ≥1 interproximal site with PPD ≥5 mm). The overall number of participants with PD was the sum of mild, moderate and severe PD.

Next, we computed for each tooth the periodontal inflamed surface area (PISA) and the periodontal epithelial surface area (PESA) for each specific participant. PISA comprised the surface area of bleeding pocket epithelium and PESA is the root surface area of that tooth, which is covered with pocket epithelium ^29,30^. PISA and PESA were calculated using a Microsoft Excel spreadsheet, in the following steps:

a. Calculation of mean CAL and gingival recession for each particular tooth;
b. Linear mean CAL and gingival recession is used to calculate PESA.
c. For each tooth, PISA is calculated through the multiplication of PESA by the proportion of sites around the tooth with bleeding on probing;
d. The sum of all individual PISA and PESA scores, in mm^2^, provides an overall area, respectively, for each participant.

### Plasma homocysteine measurement

For NHANES 2001, total Hcy in plasma was quantified by a fully automated fluorescence polarization immunoassay (FPIA)^∗ 31^. For NHANES 2002 and 2003-2004, total Hcy in plasma was measured by other FPIA^† 31–33^. Both methods employed the same reagent kit. Also, both approaches presented equivalence as the FPIA results were successfully compared to high performance liquid chromatography with fluorometric detection at 385 nm excitation and 515 nm emission ^31,33^. Hyperhomocysteinemia was defined as total Hcy in plasma >15 µmol/L ^4,5^.

### Covariates

We included sociodemographic data, including age, gender, race/ethnicity (categorized as Mexican American, Non-Hispanics White, Non-Hispanics Black, other Hispanics and other races) and education level (categorized as less than high school, complete high school or similar, higher than high school).

Smoking status was categorized as never (<100 cigarettes smoked in life and not currently smoking), former (⍰100 cigarettes in life and not currently smoking) and active smoker ((⍰100 cigarettes in life and currently smoking).

We calculated body mass index (BMI) as weight in kilograms divided by height in meters squared. For BP, systolic BP (SBP) and diastolic BP (DBP) were determined by trained and calibrated examiners. Both BP measures resulted from the average of three consecutive measurements separated by a 5-minute interval.

Blood levels data included white blood cells (WBC) Count (10^9^/L), segmented neutrophils (10^9^/L), vitamin B12 (mg/dL), hemoglobin A1c (Hba1c) (%), CRP (mg/dL), Vitamin B12 (VB12) (pg/mL) and Folate (ng/mL). VB12 and Folate were considered given its link to the Hcy biosynthesis ^1,2^.

### Data management, test methods and analysis

Databases from the NHANES 2001-2002 and 2003-2004 were uploaded and treated with r (version for Macintosh). For the purpose of periodontal diagnosis, data was exported to a spreadsheet processor^‡^ with algorithms, as previously described ^34^, to compute the periodontal status, PISA and PESA (as in Nesse et al., ^30^). After checking for data normality and homoscedasticity, measures were reported through mean (SD) for continuous variables, and a number of cases (n) and percentage (%) for categorical variables. T-test was used to compare continuous measures and chi-square test to compare categorical variables according to the periodontal status (periodontitis vs. no periodontitis).

Pearson coefficient was used to assess the correlation between the Hcy and CRP with age, BMI, periodontal clinical measures (PISA and PESA), WBC, segmented neutrophils, VB12 and Folate. A multivariate stepwise adjusted linear regression was used to model the influence of Hcy on PISA and PESA. Model variables were selected among significant variables according to the periodontal status (Table 1). An initial crude model (model 1), followed by seven progressively adjusted models were generated (model 2 - Includes adjustment for homocysteine and age; model 3 - Includes adjustment for homocysteine, age and BMI; model 4 - Includes adjustment for homocysteine, age, BMI and SBP; model 5 - Includes adjustment for homocysteine, age, BMI, SBP and WBC; model 6 - Includes adjustment for homocysteine, age, BMI, SBP, WBC and Vitamin B12; model 7 - Includes adjustment for homocysteine, age, BMI, SBP, WBC, Vitamin B12 and Folate). Multivariate stepwise adjusted linear regressions were carried out on the association of SBP and DBP with PISA or PESA (appendix S4, pp 5), and homocysteine levels and SBP or DBP (appendix S5, pp 6).

**Table 1.** Sample characteristics according to periodontal status (n=4,021).

| Variable | No periodontitis<br>(n=3,287) | Periodontitis<br>(n=734) | p-value | Overall<br>(n=4021) |
| --- | --- | --- | --- | --- |
| Age (years), mean (SD) | 38.4 (17.8) | 56.8 (15.8) | <0.001 | 41.7 (18.9) |
| Gender, % (n) |  |  |  |  |
| Males | 46.2 (1,520) | 61.6 (452) | <0.001 | 49.0 (1,972) |
| Females | 53.8 (1,767) | 38.4 (282) |  | 51.0 (2,049) |
| Race/ethnicity, % (n) |  |  |  |  |
| Mexican American | 21.0 (690) | 26.0 (191) | 0.001 | 21.9 (881) |
| Non-Hispanic White | 3.4 (111) | 4.0 (29) |  | 3.5 (140) |
| Non-Hispanic Black | 50.0 (1,643) | 41.6 (305) |  | 48.4 (1,948) |
| Other Hispanic | 21.8 (715) | 24.0 (176) |  | 22.2 (891) |
| Other race | 3.9 (128) | 4.5 (33) |  | 4.0 (161) |
| Education level, % (n) |  |  |  |  |
| < High school | 24.2 (797) | 37.9 (278) | <0.001 | 26.7 (1,075) |
| High school | 25.6 (842) | 26.8 (197) |  | 25.8 (1,039) |
| > High school | 50.1 (1,647) | 35.1 (258) |  | 47.4 (1905) |
| Smoking status, % (n) |  |  |  |  |
| Never | 63.9 (2,100) | 37.5 (275) | <0.001 | 59.1 (2,375) |
| Former | 18.3 (603) | 31.5 (231) |  | 20.7 (834) |
| Current | 17.8 (584) | 31.1 (228) |  | 20.2 (812) |
| BMI (kg/m<sup>2</sup>), mean (SD) | 27.7 (7.0) | 28.5 (7.2) | 0.011 | 27.9 (7.1) |
| Homocysteine (μmol/L), mean (SD) | 8.2 (3.9) | 9.8 (4.0) | <0.001 | 8.5 (4.0) |
| Homocysteine elevated level category, % (n) | 2.8 (92) | 6.1 (45) | <0.001 | 3.4 (137) |
| CRP (mg/dL), mean (SD) | 0.4 (0.8) | 0.5 (0.8) | <0.001 | 0.4 (0.8) |
| Chronic medical conditions, mean (SD) | 0.8 (1.5) | 1.4 (2.7) | <0.001 | 0.9 (1.8) |
| Diabetes, n (%) | 5.0 (164) | 14.7 (108) | <0.001 | 6.8 (272) |
| Hypertension, n (%) | 12.1 (399) | 29.4 (216) | <0.001 | 15.3 (615) |
| Mean SBP (mmHg), mean (SD) | 120.0 (17.6) | 132.0 (21.2) | <0.001 | 122.2 (18.9) |
| Mean DBP (mmHg), mean (SD) | 69.4 (11.7) | 72.0 (11.9) | <0.001 | 69.8 (11.8) |
| PISA (mm<sup>2</sup>), mean (SD) | 0.8 (7.9) | 13.5 (38.4) | <0.001 | 3.1 (18.5) |
| PESA (mm<sup>2</sup>), mean (SD) | 17.3 (51.4) | 118.0 (107.0) | <0.001 | 35.6 (75.7) |
| Vitamin B12 (pg/mL), mean (SD) | 550.0 (1,120.0) | 554.0 (521.0) | 0.880 | 550.9 (1,037.1) |
| Folate (ng/mL), mean (SD) | 12.6 (7.8) | 14.6 (26.3) | 0.040 | 12.9 (13.3) |

The mediating effect of PISA and PESA in the association of Hcy with SBP and DBP was carried out using the r package ‘lavaan’. Mediation analysis was done through the establishment of three pathways (a, b and c) (appendix S2, pp 3). Total effect (c-path) evaluated the relationship between the exposure (Hcy) and the outcomes of interest (SBP or DBP). The a-path assessed the direct effect of the exposure (Hcy) on the mediators of interest (PISA or PESA). The b-path measured the mediators (PISA or PESA) direct effect on the outcomes of interest (SBP or DBP). The mediation effect was computed through multiplying a-path with b-path. The proportion of the mediated effect was calculated using the following formula: (total effect - direct effect) x 100. All mediation models have been adjusted for sociodemographic variables (age, gender, race, education), smoking habit, BMI, systemic status (number of chronic medical conditions, hypertension, diabetes, hba1c), vitamin B12 and folate. A value of p<0.05 was considered significant.

## Results

### Characteristics of the Study Sample

With a primary sample of 21,161 participants, 17,140 were excluded based on the defined the inclusion criteria, thus, resulting in a final sample of 4,021 patients (appendix S3, pp 4). The overall characteristics of these participants are shown in Table 1. The mean age of the participants was 41.7 (±18.9) years, with balanced proportion between males and females. Participants were mostly non-hispanic black participants (48.4%), higher education level (47.4%) and self-reported as never smoker (59.1%). Also, this sample had an average body mass index (BMI) of 27.9 (±7.1) kg/m^2^. A total of 734 participants were diagnosed as periodontitis cases (18.3%). Periodontitis cases presented significantly differences regarding mean age, gender, race/ethnicity, education level, smoking status, Hcy mean levels, Hcy category levels, chronic medical conditions, diabetes, hypertension, BP (Systolic BP and Diastolic BP), CRP, periodontal inflamed surface area (PISA) and periodontal epithelial surface area (PESA) (p<0.001). Mean BMI (p=0.011) and folate (p=0.040) were also different among periodontal groups, however, Vitamin B12 was not (p=0.880).

### Correlation estimates of Hcy compared to CRP

To understand if Hcy and CRP displayed alike behavior with similar variables, we investigated the correlation of Homocysteine and CRP with relevant continuous variables (Table 2). In the Hcy levels, age had a moderate correlation effect (p<0.001), while SBP, DBP, PISA and PESA had a mild correlation effect (p<0.001). Furthermore, we confirmed the association of Hcy levels with Vitamin B12 and folate (p<0.001). In the overall sample, CRP had a significantly mild correlation with BMI (p<0.001), WBC (p<0.001), Segmented Neutrophils (p<0.001) and Age (p<0.05). The correlations between PISA and PESA with SBP and DBP were graphically displayed (p<0.01) (Figure 2).

**Table 2.** Correlation of CRP and Homocysteine with sociodemographic and clinical variables (n=4,021).

| Variable | CRP (mg/dL) | Homocysteine (μmol/L) |
| --- | --- | --- |
| Age (years) | 0.036* | 0.326** |
| Homocysteine (μmol/L) | -0.019 | - |
| BMI (g/m<sup>2</sup>) | 0.215** | 0.003 |
| SBP (mmHg) | 0.037* | 0.274** |
| DBP (mmHg) | 0.015 | 0.124** |
| PISA (mm<sup>2</sup>) | 0.030 | 0.054** |
| PESA (mm<sup>2</sup>) | 0.031 | 0.177** |
| WBC (10<sup>-9</sup>) | 0.154** | -0.026 |
| Segmented Neutrophils (10<sup>-9</sup>) | 0.143** | -0.035* |
| Vitamin B12, (pg/mL) | 0.006 | -0.058** |
| Folate, (ng/mL) | -0.011 | -0.045** |

Pearson correlation, \* p < 0.05, \*\* p < 0.001.
BMI – Body Mass Index; CRP – C-reactive Protein; DBP – Diastolic Blood Pressure; PISA – Periodontal Inflamed Surface Area; PESA – Periodontal Epithelial Surface Area; SBP – Systolic Blood Pressure; WBC – White blood cells counts.

**Figure 1.**
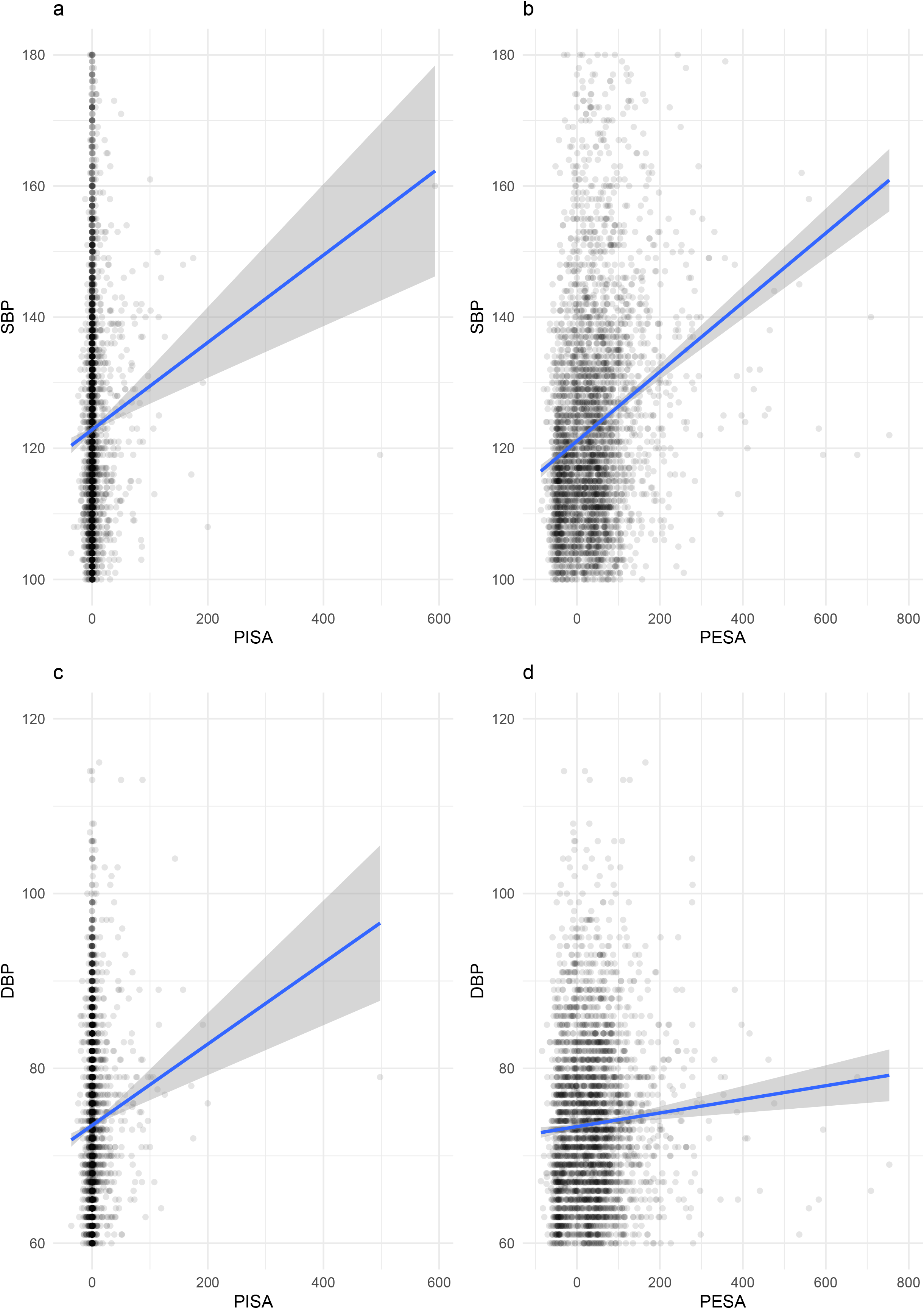
Scatterplots displaying the correlation between PISA with SBP (a) and DBP (c), and PESA with SBP (b) and DBP (d).

Next, we examined the linear relationship of Hcy with PISA and PESA in crude (model 1) and adjusted models (models 2-8) (Table 3). Linear regression models confirmed that PISA is significantly associated with Hcy (p<0.001) however, that not occurred in the adjusted models. While for PESA, both for the crude and adjusted models, it was significantly associated with Hcy (p<0.01). The linear association of SBP and DBP with PISA and PESA was also carried out (appendix S4, pp 5), with DBP being associated with PISA in all adjusted models. The linear association of Hcy with SBP and DBP was found to be significant in all adjusted models (appendix S5, pp 6).

**Table 3.** Crude and adjusted linear regression models of homocysteine levels and PISA or PESA for the overall sample with the respective B coefficient and standard error (SE) (n=4,021).

|  | Homocysteine |  |
| --- | --- | --- |
|  | PISA | PESA |
| Model 1 | 0.012** (0.003) | 0.009** (0.001) |
| Model 2 | 0.005 (0.003) | 0.002* (0.001) |
| Model 3 | 0.005 (0.003) | 0.002* (0.001) |
| Model 4 | 0.004 (0.003) | 0.002* (0.001) |
| Model 5 | 0.004 (0.003) | 0.002* (0.001) |
| Model 6 | 0.004 (0.003) | 0.002* (0.001) |
| Model 7 | 0.003 (0.003) | 0.002* (0.001) |

Values are presented as B coefficient (SE).

### Mediation analyses

There was evidence that the association between Hcy with BP (SBP and DBP) were mediated by PISA and PESA. Mediation analyses demonstrated that PISA was estimated to mediate 17.39% and 16.33% of the total association between Hcy with SBP and DBP, respectively (p<0.001) (Table 4). Furthermore, PESA was estimated to mediate 0.89% and 47.20% of the total association between Hcy with SBP and DBP, respectively (p<0.001) (Table 5).

**Table 4.** Mediation analysis of the effects of PISA and PESA on the association of homocysteine levels with systolic blood pressure (n = 4,021).

| Exposure: Homocysteine / Outcome: SBP |  | Mediator: PISA |  |
| --- | --- | --- | --- |
| Effect | Estimate | SE | p-value |
| a (exposure → mediator) | 0.156 | 0.076 | 0.041 |
| b (mediator → outcome) | 0.076 | 0.017 | <0.001 |
| c (total effect) | 0.069 | 0.006 | <0.001 |
| c' (direct effect) | 0.012 | 0.003 | <0.001 |
| ab (mediated effect) | 0.012 | 0.006 | <0.001 |
| ab/c (PISA percentage mediated) = 17.4% |  |  |  |
| Exposure: Homocysteine / Outcome: SBP |  | Mediator: PESA |  |
| Effect | Estimate | SE | p-value |
| a (exposure → mediator) | 0.006 | 0.001 | <0.001 |
| b (mediator → outcome) | 1.176 | 0.074 | <0.001 |
| c (total effect) | 0.900 | 0.063 | <0.001 |
| c' (direct effect) | 0.892 | 0.063 | <0.001 |
| ab (mediated effect) | 0.008 | 0.001 | <0.001 |
| ab/c (PESA percentage mediated) = 0.9% |  |  |  |

Models adjusted for sociodemographic variables (age, gender, race, education), smoking habit, BMI, systemic status (number of chronic medical conditions, hypertension, diabetes, hba1c), vitamin B12 and folate. Abbreviations: PISA – Periodontal Inflamed Surface Area; PESA – Periodontal Epithelial Surface Area; SBP – Systolic Blood Pressure.

**Table 5.**
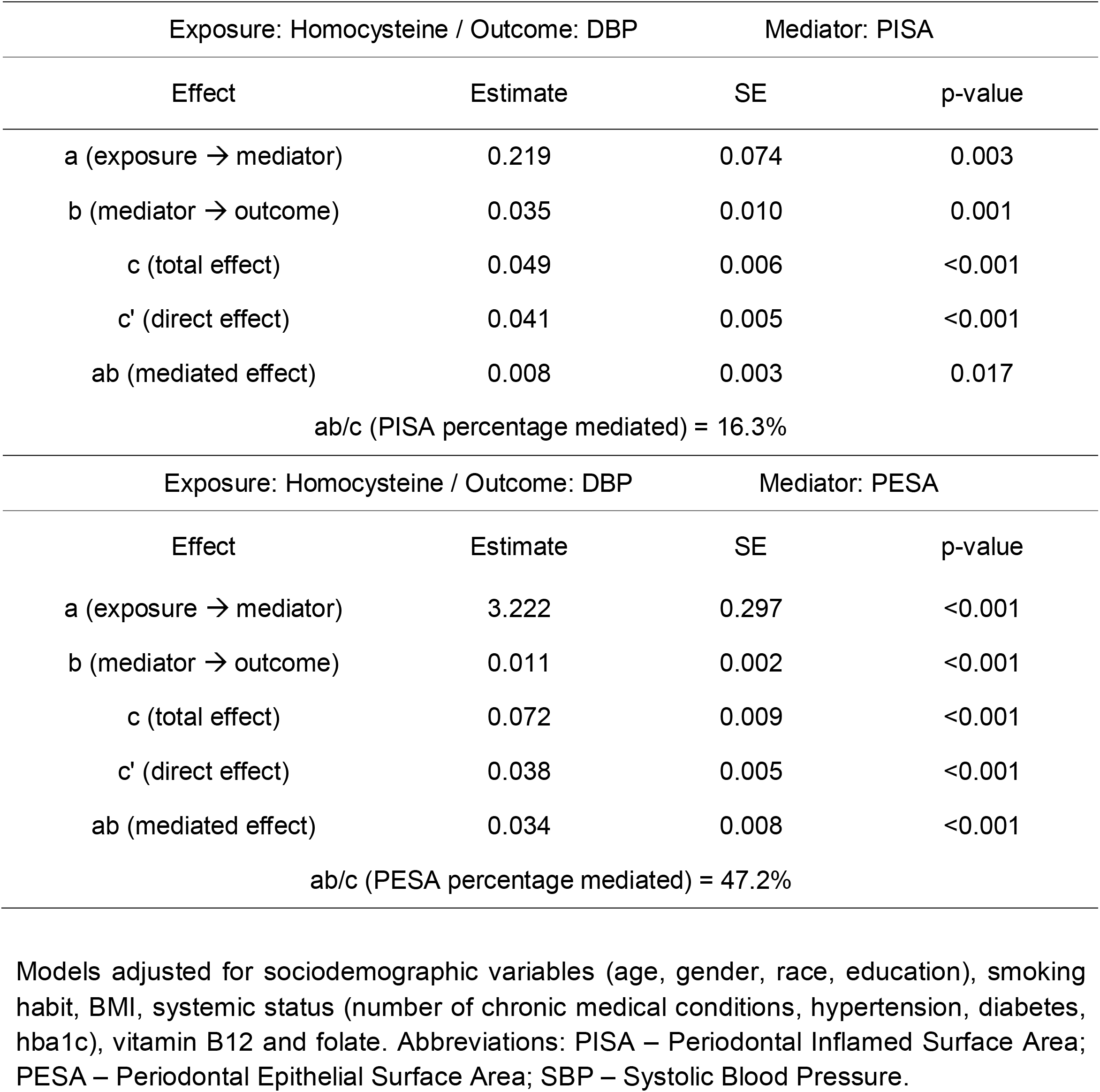
Mediation analysis of the effects of PISA and PESA on the association of homocysteine levels with diastolic blood pressure (n = 4,021).

## Discussion

In this exploratory study, we found periodontitis cases to present higher Hcy levels, and this association was significantly associated even when adjusted for multiple confounding variables. These findings, therefore, support an association between Hcy and periodontitis, and periodontitis measures (PISA and PESA) as mediators in the link between Hcy and BP.

These results, in our view, are unique because, this large-based study with a comprehensive set of variables support this association and strengthen the role of periodontitis in the association of Hcy with high blood pressure. Seemingly, this association relies more on the destruction of periodontal tissues (PESA) rather than inflamed surface area (PISA).

In fact, Hcy was initially considered in Periodontal Medicine as a candidate in the link of periodontal and cardiovascular conditions ^20^. Comprehensively, Hcy inhibits nitric oxide in the endothelium affecting its function and leads to excessive reactive oxygen radicals in the myocardium ^18,19^. The high level of homocysteine in serum makes a person more prone to endothelial cell injury, and is collectively seen as an inflammatory reaction in the blood vessels. In a recent systematic review, periodontitis was proposed to affect vascular function and triggering hypertension through boosted systemic inflammation and bacteriemia, instigating vascular damage (by increased cytokines, immune cells, nitric oxide and reactive oxygen species) ^26^. Later, studies with small samples reported this association ^23,24^. Notwithstanding, our study may stand-out from the others by the considerable increase in participants.

In this sense, and given the cross-sectional nature of this study (also discussed later), we cannot discern whether this is a unilateral (and triggered by Hcy or periodontitis) or bilateral nature. On the one hand, the increased inflammatory burden driven by untreated periodontitis is consistently proven to increase the risk towards cardiovascular disease ^35^. On the other hand, indirect evidence from a clinical trial demonstrated that adjunctive folate during periodontal treatment reduced plasma homocysteine, yet with unknown long-term clinical consequences (i.e., reduced risk of high blood pressure) ^36^.

Lastly, the fact that PESA was more significant as mediator in the Hcy-BP axis than PISA possibly points to an unknown impact of the periodontium tissues destruction. The bacterial role related to methionine pathways relies on the production of volatile sulfur compounds by periodontal bacteria, causing the commonly detected malodor in periodontal patients ^37–39^. At this stage, this results only highlight a possible connection point, and the reader must bear in mind that they are solely hypothetical. Though, the invasion of periodontal bacteria through the ulcerated epithelium into the blood stream has been shown, with prospective hostile systemic consequences ^40,41^.

### Strengths and Limitations

The present cross-sectional study has specific fallouts worth examining. The observational nature of NHANES precluded any inference of causality or temporal link. Furthermore, information regarding systemic inflammation was limited as CRP levels was the only recognized and accepted hallmark available, thus limiting the validity of these results. For this reason, data on other important markers of inflammation, such as cytokines (TNF-a and interleukins) might be of interest to study in future studies.

When exploring the association and mediated effect, these results should be interpreted cautiously, however the causal association has been demonstrated with recent prospective studies and randomized intervention trials ^20–25,36^. This sample is based on two consecutive NHANES waves, and are representative of the American population. The number of covariates included in our analyses were comprehensive.

The periodontal assessment was made by calibrated examiners yet a partial-mouth protocol has been taken place, which may contribute to less accurate and precise results ^34,42^. Despite this, our estimates revealed very interesting values of both correlation and mediation.

In conclusion, Hcy serum levels are associated with the periodontal status. Hcy levels demonstrated more relationship with PISA and PESA, BP and age. Compared to CRP, Hcy showed different associations. Besides, PISA and PESA might play a mediation effect on the link between Hcy and BP.

## Acknowledgments

We acknowledge the NHANES investigators and personnel.

## Authors contribution statement

The authors’ responsibilities were as follows— JB and VM designed the research; JB conducted the research; JB, VM and LP analyzed the data and performed the statistical analysis; JB, VM, YL, LP, JJM wrote the manuscript; JB has primary responsibility for final content; and all authors contributed to critically reviewing the manuscript and read and approved the final manuscript. All authors declare no conflicts of interest.

## FOOTNOTES

∗ Abbott Homocysteine IMX (HCY) assay

† Abbott Diagnostics (Abbott AxSym analyzer, Abbott®)

‡ Microsoft Office Excel

