## Supplementary files for "Periodontal inflammation mediates the link between homocysteine and high blood pressure"

**Appendix S1.** STROBE Statement—Checklist of items that should be included in reports of ***case-control studies***

|  | **Item No** | **Recommendation** | **Page** |
| --- | --- | --- | --- |
| **Title and abstract** | 1 | (*a*) Indicate the study’s design with a commonly used term in the title or the abstract | 1 |
|  |  | (*b*) Provide in the abstract an informative and balanced summary of what was done and what was found | 2 |
| **Introduction** | | |  |
| Background/rationale | 2 | Explain the scientific background and rationale for the investigation being reported | 4 |
| Objectives | 3 | State specific objectives, including any prespecified hypotheses | 5 |
| **Methods** | | |  |
| Study design | 4 | Present key elements of study design early in the paper | 5 |
| Setting | 5 | Describe the setting, locations, and relevant dates, including periods of recruitment, exposure, follow-up, and data collection | 5 |
| Participants | 6 | (*a*) Give the eligibility criteria, and the sources and methods of case ascertainment and control selection. Give the rationale for the choice of cases and controls | 5 |
|  |  | (*b*) For matched studies, give matching criteria and the number of controls per case |  |
| Variables | 7 | Clearly define all outcomes, exposures, predictors, potential confounders, and effect modifiers. Give diagnostic criteria, if applicable | 5 |
| Data sources/ measurement | 8* | For each variable of interest, give sources of data and details of methods of assessment (measurement). Describe comparability of assessment methods if there is more than one group |  |
| Bias | 9 | Describe any efforts to address potential sources of bias |  |
| Study size | 10 | Explain how the study size was arrived at | 5 |
| Quantitative variables | 11 | Explain how quantitative variables were handled in the analyses. If applicable, describe which groupings were chosen and why | 5-9 |
| Statistical methods | 12 | (*a*) Describe all statistical methods, including those used to control for confounding | 9 |
|  |  | (*b*) Describe any methods used to examine subgroups and interactions |  |
|  |  | (*c*) Explain how missing data were addressed |  |
|  |  | (*d*) If applicable, explain how matching of cases and controls was addressed |  |
|  |  | (*e*) Describe any sensitivity analyses |  |
| **Results** | | |  |
| Participants | 13* | (a) Report numbers of individuals at each stage of study—eg numbers potentially eligible, examined for eligibility, confirmed eligible, included in the study, completing follow-up, and analysed | 10 |
|  |  | (b) Give reasons for non-participation at each stage | 10 |
|  |  | (c) Consider use of a flow diagram | 10 |
| Descriptive data | 14* | (a) Give characteristics of study participants (eg demographic, clinical, social) and information on exposures and potential confounders | 11 |
|  |  | (b) Indicate number of participants with missing data for each variable of interest | NA |
| Outcome data | 15* | Report numbers in each exposure category, or summary measures of exposure | 11 |
| Main results | 16 | (*a*) Give unadjusted estimates and, if applicable, confounder-adjusted estimates and their precision (eg, 95% confidence interval). Make clear which confounders were adjusted for and why they were included | 11 |
|  |  | (*b*) Report category boundaries when continuous variables were categorized | 11 |
|  |  | (*c*) If relevant, consider translating estimates of relative risk into absolute risk for a meaningful time period | 11 |

**Appendix S2.** Path diagram of the mediation analysis models.

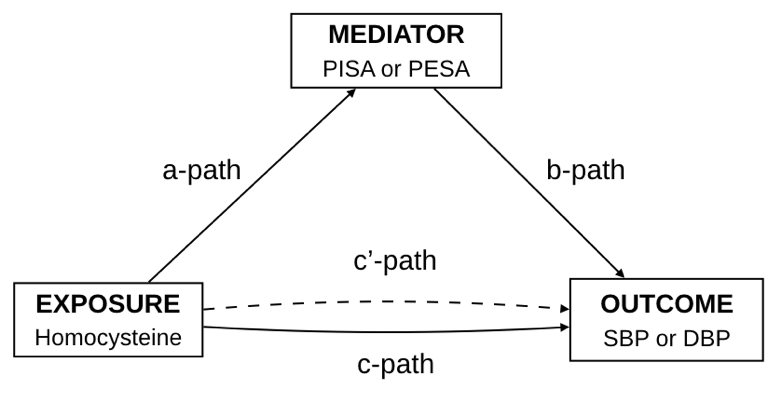

**Appendix S3.** Flowchart of participants.

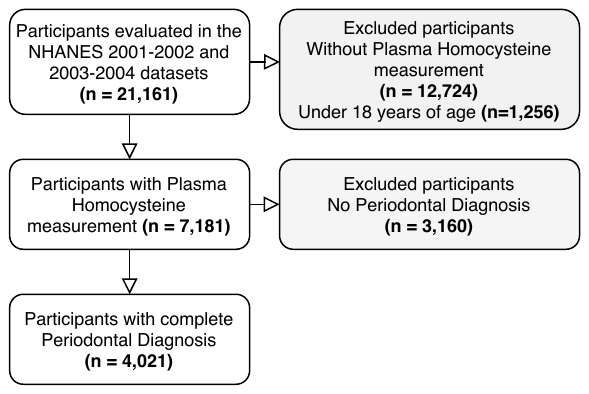

**Appendix S4.** Crude and adjusted linear regression models of SBP and DBP with PISA or PESA for the overall sample with the respective B coefficient and standard error (SE) (n=4,021).

|  | SBP | | DBP | |
| --- | --- | --- | --- | --- |
|  | PISA | PESA | PISA | PESA |
| Model 1 | 0.085*** (0.016) | 0.063 (0.003)*** | 0.039 (0.010)*** | 0.014 (0.002)*** |
| Model 2 | 0.028* (0.013) | -0.001 (0.004) | 0.024 (0.010)* | -0.003 (0.003) |
| Model 3 | 0.025 (0.013) | -0.001 (0.004) | 0.022 (0.010)* | -0.002 (0.003) |
| Model 4 | 0.023 (0.013) | -0.001 (0.004) | 0.021 (0.010)* | -0.002 (0.003) |
| Model 5 | 0.023 (0.013) | -0.001 (0.004) | 0.022 (0.010)* | -0.002 (0.003) |
| Model 6 | 0.023 (0.013) | -0.001 (0.004) | 0.022 (0.010)* | -0.002 (0.003) |
| Model 7 | 0.023 (0.013) | -0.001 (0.004) | 0.022 (0.010)* | -0.002 (0.003) |

Values are presented as B coefficient (SE).

Model 1 - Unadjusted model; Model 2 - Includes adjustment for age; Model 3 - Includes adjustment for age and BMI; Model 4 - Includes adjustment for age, BMI and Homocysteine; Model 5 - Includes adjustment for age, BMI, Homocysteine and WBC; Model 6 - Includes adjustment for age, BMI, Homocysteine, WBC and Vitamin B12; Model 7 - Includes adjustment for age, BMI, Homocysteine, WBC, Vitamin B12 and Folate. * p < 0.05; ** p < 0.01; *** p < 0.001.

**Appendix S5.** Crude and adjusted linear regression models of homocysteine levels and SBP or DBP for the overall sample with the respective B coefficient and standard error (SE) (n=4,021).

|  | Homocysteine | |
| --- | --- | --- |
|  | SBP | DBP |
| Model 1 | 0.058*** (0.003) | 0.042*** (0.005) |
| Model 2 | 0.028*** (0.004) | 0.018*** (0.005) |
| Model 3 | 0.029*** (0.004) | 0.020*** (0.005) |
| Model 4 | 0.029*** (0.004) | 0.019*** (0.005) |
| Model 5 | 0.029*** (0.004) | 0.020*** (0.005) |
| Model 6 | 0.029*** (0.004) | 0.020*** (0.005) |
| Model 7 | 0.029*** (0.004) | 0.020*** (0.005) |
| Model 8 | 0.029*** (0.004) | 0.019*** (0.005) |

Values are presented as B coefficient (SE).

Model 1 - Unadjusted model; Model 2 - Includes adjustment for age; Model 3 - Includes adjustment for age and BMI; Model 4 - Includes adjustment for age, BMI and PISA; Model 5 - Includes adjustment for age, BMI, PISA and PESA; Model 6 - Includes adjustment for age, BMI, PISA, PESA and WBC; Model 7 - Includes adjustment for age, BMI, PISA, PESA, WBC, and Vitamin B12; Model 8 - Includes adjustment for age, BMI, PISA, PESA, WBC, Vitamin B12 and Folate. * p < 0.05; ** p < 0.01; *** p < 0.001.
